# Hospital Waste Management: An Overview of the Situation in Health Districts outside Yaounde in the Centre Region, Cameroon

**DOI:** 10.1101/2024.12.08.24318669

**Authors:** Michel Franck Edzamba, Fabrice Zobel Lekeumo Cheuyem, Paule Sandra Djomo Sime, Fernande Eliane Nyangon Minlo, Bertille Ngon Sani, Florence Kissougle Nkongo, Tatiana Mossus

**Affiliations:** Department of Public Health, Faculty of Medicine and Biomedical Sciences, The University of Yaounde I, Yaounde, Cameroon; Regional Delegation of Public Health for the Centre Region, Ministry of Public Health, Yaounde, Cameroon

**Keywords:** Health facility, waste management, Centre Region, Cameroon

## Abstract

**Background:** Hospital waste management is an important process that must be handled with care. This is because of the potential risks it represents for human health and the environment. A good understanding of the processes and legislation involved is an important asset, especially in developing countries. In order to assess the level of application of regulations, this study describes the quality of hospital waste management in health districts outside Yaounde in the Centre region.

**Methodology:** A descriptive cross-sectional study was conducted in public health facilities of 24 Health Districts in the Centre Region from February 1 to March 31, 2024. Sampling was consecutive and non-exhaustive. Data were collected using a Google form from health facility managers and/or staff in charge of hospital hygiene. The data was coded and analyzed using R Statistics version 4.3.1.

**Results:** A total of 61 participants coming from 20 Health Districts of the Central Region were enrolled in the study. With a sex ratio (M/F) of 0.9, most of the participants were paramedical staff. Less than 10% of the participants were Medical-sanitary technician. Most of the respondents reported that waste was sorted at source. However, nearly half (45%) carried out regular checks at this stage. More than two-thirds of responded (78%) reported that household waste was packed in buckets and black bags. More than half of the infectious waste was packaged in bin buckets (57%). More than one-third (39%) of the waste was collected daily. The vast majority (93%) reported that they disposed of waste on site. Nearly 5% reported disposing of waste in a public landfill. A small proportion (7.3%) of health facilities had an incinerator.

**Conclusion:** Despite the efforts made, there are still significant shortcomings in the hospital waste management strategy of the health facilities visited. Stakeholders need technical, financial and material support if they are to meet international standards.

## Introduction

Healthcare activities generate a growing amount of hospital waste, which is increasing in quantity and diversity worldwide [1]. It is estimated that medical waste represents approximately 1 to 2% of all municipal waste generated [2]. Some healthcare waste poses no particular risk and can be handled in the same way as household waste. Others, however, pose infectious risks [3]. The healthcare waste is defined as all waste generated in healthcare settings, research centers and laboratories in connection with medical procedures, as well as waste from minor and dispersed sources, including waste generated in the context of healthcare carried out at home, such as home dialysis, insulin self-administration and recovery care [4]. In the hospital environment, rigorous management of all healthcare waste is a key component to improve the quality and safety of care and to prevent adverse events in the healthcare environment, in particular the prevention of healthcare-associated infections [5].

Systems for the storage, handling, treatment and disposal of medical (healthcare) waste are well developed in the 21st century. However, in many low-and middle-income countries, these systems, resources and expertise are lacking, to such an extent that medical waste could pose a serious threat to the health, safety and lives of millions of healthcare workers and waste handlers who frequently come into contact with this category of material [6]. It is estimates that approximately 85% of healthcare waste is non-hazardous (comparable to household waste), while 10% is infectious (cultures and stocks of infectious agents, waste from infected patients, waste contaminated with blood and blood derivatives, discarded diagnostic specimens, infected animals from laboratories, and contaminated materials and equipment) and anatomical waste (identifiable body parts and animal cadavers) and the remaining 5% is hazardous healthcare waste (chemical, radioactive) [7]. Many developed countries apply strict guidelines for the segregation, storage and transport of medical waste [8]. Developing countries, on the other hand, are faced with limited resources when it comes to effectively manage hospital waste [9]. In this case, poor sanitation practices can lead to the mixing of hazardous waste with general waste, which can exacerbate the waste management problem by increasing the cost of treatment and disposal [10]. In addition, poor nutrition, inadequate healthcare and lack of immunization can increase the public’s susceptibility to infection from untreated medical waste [9].

In developing countries, there is the added hazard of searching for landfill and manually sorting collected waste on at leaves healthcare settings. Similarly, the methods used to manage healthcare waste can themselves pose a health risk if the various stages in the management process are not carried out appropriately [11]. Poor risk management can endanger healthcare workers, patients and their families, and the population as a whole, not to mention the underlying environmental disasters.

In Cameroon, there are a number of texts governing hospital waste management, but they do not take into account all the types of waste that can be generated by a healthcare settings [12,13]. With the aim of assessing the level of application of the regulations, this study evaluates the hospital waste management system in the health districts outside Yaounde in the Centre region

## Methodology

### Study design and period

A descriptive, cross-sectional study was conducted from February to March 2024 in Health Districts outside Yaounde in the Central Region. It was a mixed-methods study, including both quantitative and qualitative data.

### Settings

The Centre Health Region covers 74,054 km2. The population in 2023 is estimated at 5,266,096 inhabitants. It is the second most populous in Cameroon after the Far-North. It accounts for around 18% of the total population, with a density of 73 inhabitants per km^2^. It is subdivided into 32 districts, 28 of which are outside the city of Yaounde, the political capital of Cameroon (Figure 1).

**Fig 1.**
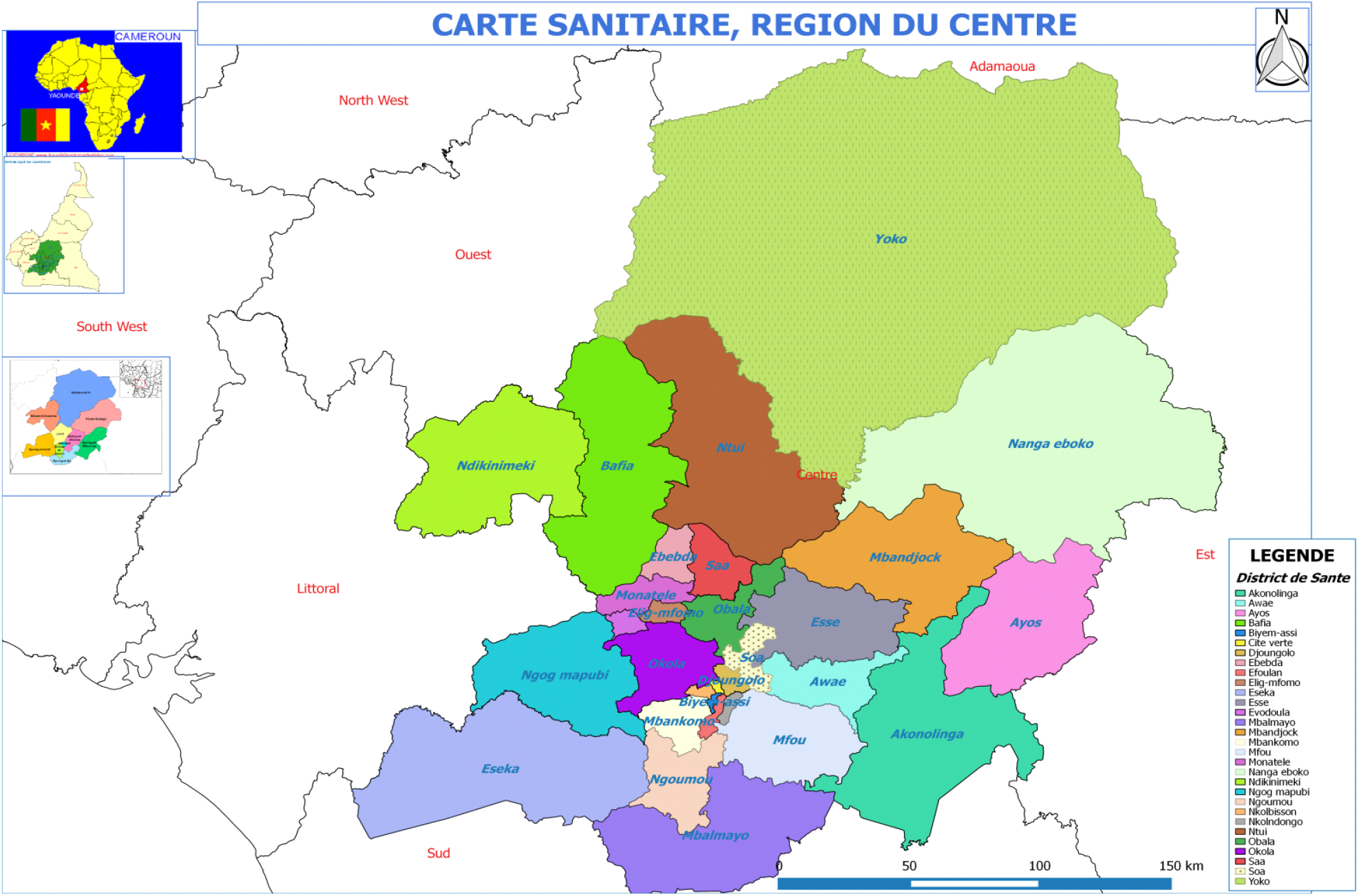
Map of the Centre Health Region (*source: Ministry of public health, Cameroon 2019)*

It comprises 287 Health Areas, 11 first and second category Hospitals, 01 Regional Annex Hospital (HRA), 30 District Hospitals, 29 Subdivisional Medical Centers for a total of 1431 health facilities, 469 of which are public, 850 secular and 112 denominational.

### Study participants and selection criteria

Health facility managers or hospital hygiene staff who consented to take part in the study were included. We used a consecutive, non-exhaustive sampling to recruit study participants.

### Data collection tools and procedures

Data were collected using a pre-designed Google Forms questionnaire, which was sent to the various managers after they had received an information letter. The questionnaire collected socio-professional information, institutional aspects and the hospital waste management indicators (sorting-collection-storage-transport-treatment-disposal). Qualitative data were collected from all participants through interviews.

### Data processing and analysis

The data collected was reviewed and checked for completeness before being analysis. Questionnaires were stored in a secure location accessible only to the principal investigator.

Descriptive statistics were carried out using R Statistics 4.3.1. Content analysis techniques was used to identify suggestions for improving the quality of hospital waste management.

## Results

A total of 61 participants coming from 20 Health Districts (83% Health Districts located outside Yaounde) of the Central Region were enrolled in the study. The sex ratio (M/F) was 0.9. Paramedical staff were the most represented. Less than 10% were medical-sanitation technician. Over 30% had more than 5 years’ experience in hospital waste management (Table 1).

**Table 1.** Socio-professional characteristics of participants, Centre Region, March 2024 (*n*=61)

| Variable | Count(n) | Frequency (%) |
| --- | --- | --- |
| **Gender** |  |  |
| Female | 32 | 52.5 |
| Male | 29 | 47.5 |
| **Professional status** |  |  |
| Maintenance agent | 7 | 11.5 |
| Care assistant | 14 | 23.0 |
| Medical-sanitary technician | 4 | 6.6 |
| Nurse | 26 | 42.6 |
| Physician | 8 | 13.1 |
| Mid-wife | 2 | 3.3 |
| **Position** |  |  |
| Health facility manager | 13 | 21.3 |
| Hygiene manager | 48 | 78.7 |
| **Experience in waste management (year)** |  |  |
| <1 | 13 | 21.3 |
| 2-4 | 28 | 45.9 |
| 5+ | 20 | 32.8 |

Most of the respondents stated that they sort their waste at source. However, almost half (46%) regularly check this stage (Table 2).

**Table 2.** Characteristics of waste sorting and packaging, Centre Region, March 2024 (*n*=61)

| Variable | Count(n) | Frequency (%) |
| --- | --- | --- |
| Waste separation | 59 | 96.7 |
| Existence of a sharp's container | 47 | 77.0 |
| Regular checks on waste sorting and packaging | 28 | 45.9 |
| **Inspection frequency** |  |  |
| Twice a month | 3 | 4.9 |
| Every week | 8 | 13.1 |
| Monthly | 7 | 11.5 |
| Quarterly | 6 | 9.8 |
| None | 37 | 60.6 |

**Table 3.** Characteristics of waste collection, storage and transport facilities, Centre Region, March 2024 (n=61)

| Variable | Count (n) | Frequency (%) |
| --- | --- | --- |
| **Collection frequency** |  |  |
| Twice a month | 2 | 3.3 |
| Monthly | 6 | 9.8 |
| Quarter | 1 | 1.6 |
| Weekly | 24 | 39.3 |
| None | 4 | 6.6 |
| Daily | 24 | 39.3 |
| **Storage times in the hospital** |  |  |
| 24 hours | 4 | 6.6 |
| 48 hours | 14 | 22.9 |
| 72 hours | 2 | 3.3 |
| 1 week | 16 | 26.2 |
| None | 17 | 27.8 |
| Other | 8 | 13.1 |
| **Existence of a transport vehicle** |  |  |
| Yes | 4 | 6.6 |
| No | 57 | 93.4 |
| **Authorized transport** |  |  |
| Yes | 3 | 4.9 |
| No | 1 | 1.6 |

Most of the household waste was packed either in cardboard boxes, in black bag or in in garbage buckets (90%) (Figure 2).

**Fig 2.**
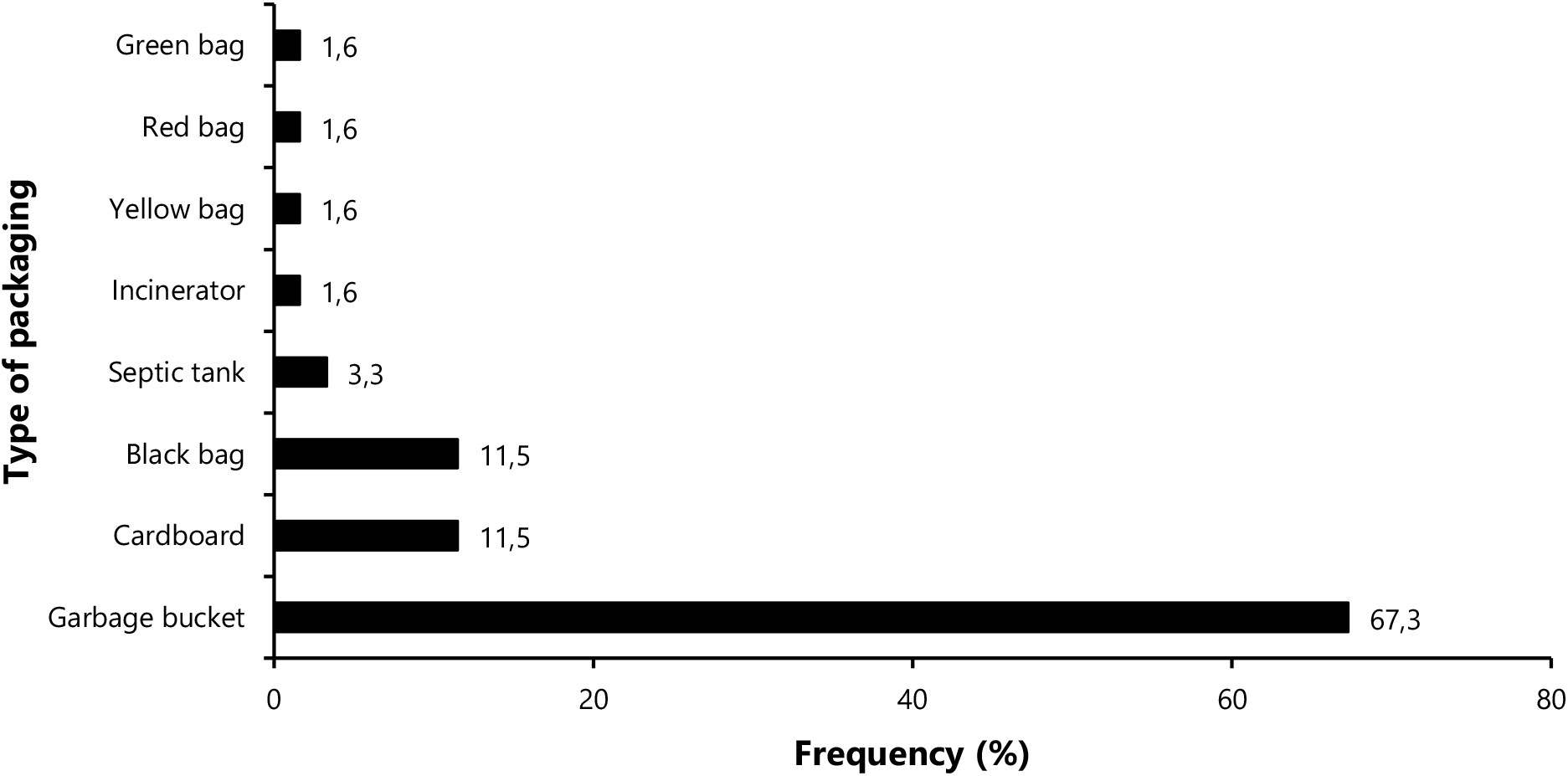
Household waste packaging by material type, Centre Region, March 2024 (*n*=61)A small proportion (5%) of soft infectious waste was packaged in yellow bags. It was found that almost 60% of soft infectious waste was placed in garbage buckets (Figure 3).

**Fig 3.**
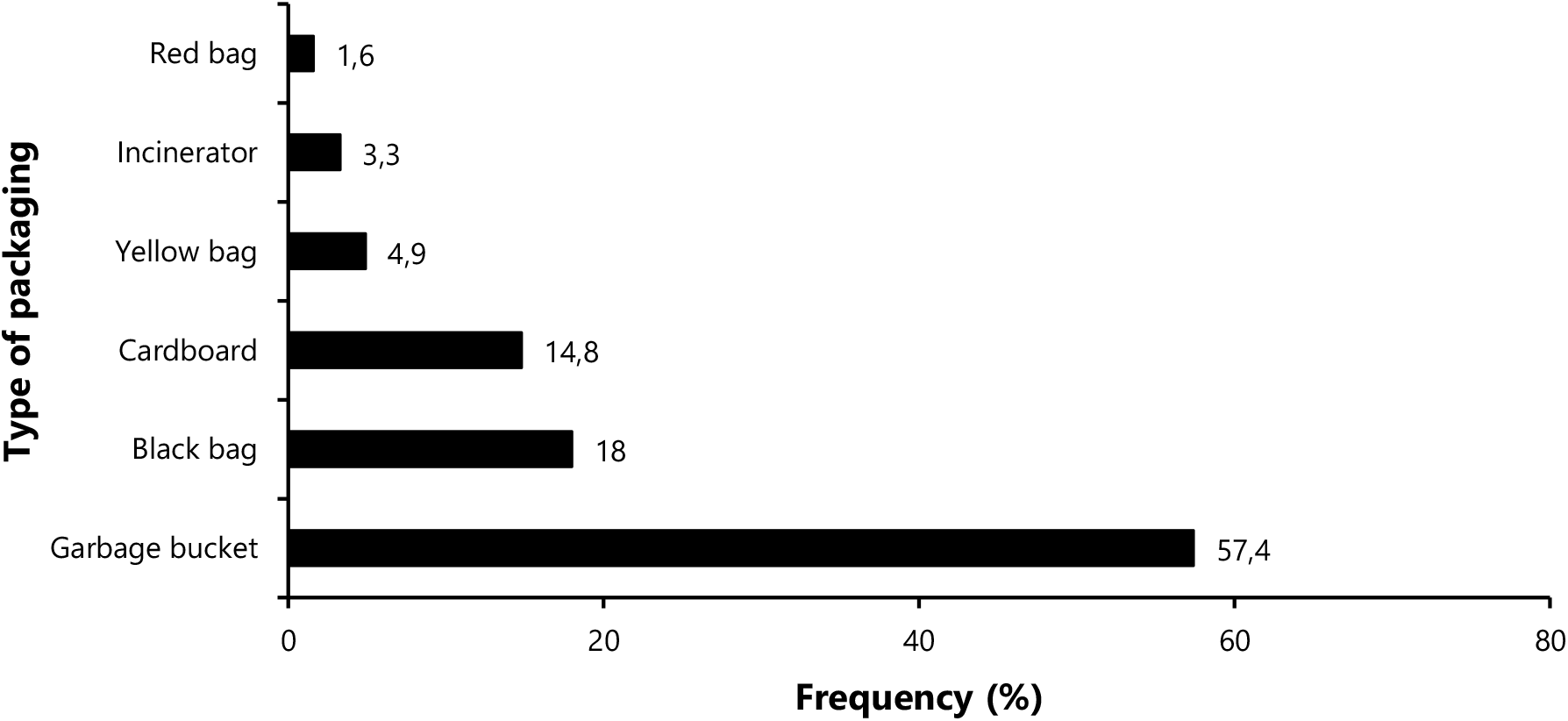
Packaging of soft infectious waste according to equipment used, Center Region, March 2024 (n=61)Almost 7% of sharps waste was found in colored bags (red and yellow), and most of these were collected in safety boxes (77.7%). Around 11% of sharps waste was packaged in cardboard boxes (Figure 4).

**Fig 4.**
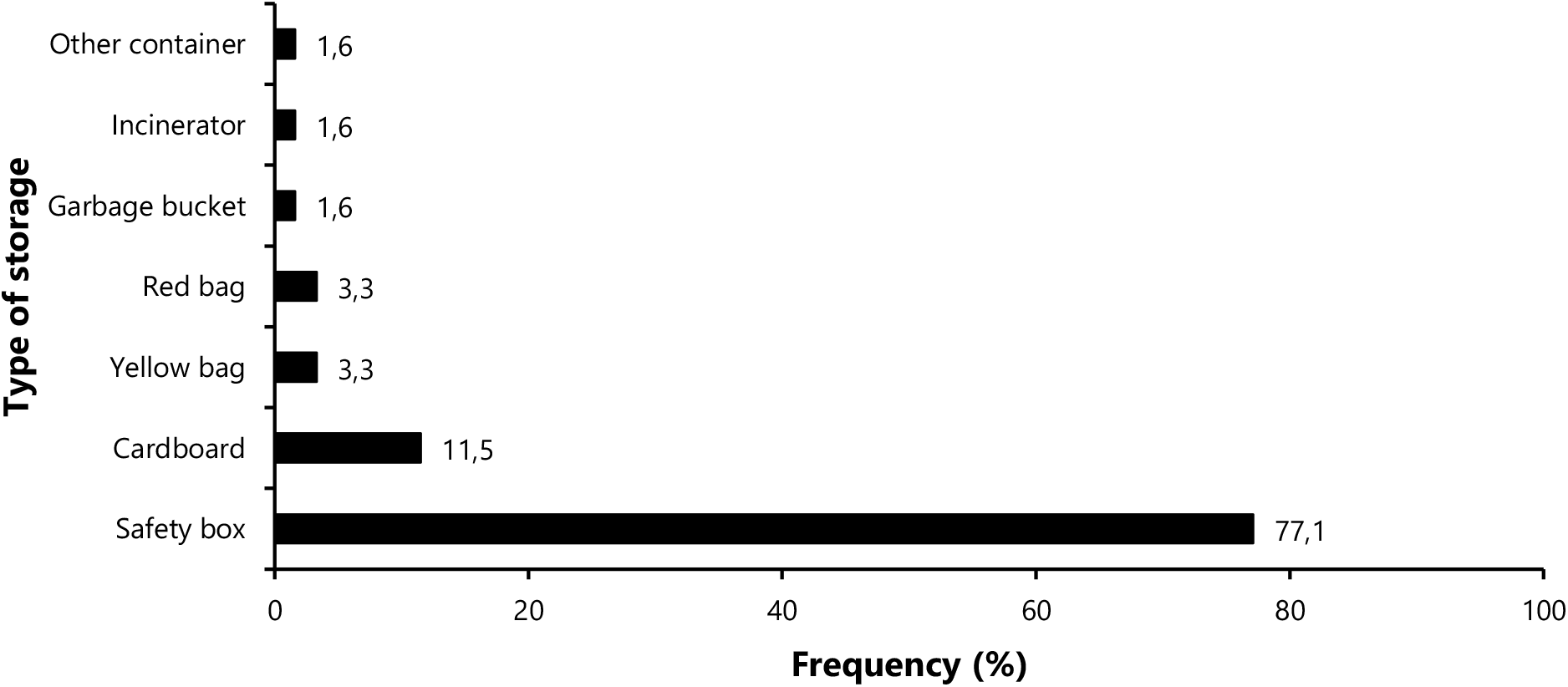
Packaging of sharps waste, Centre Region, March 2024 (n=61)Less than half (39%) of the waste was collected daily with no storage time (27%) before transport (Table 3).

Most of the respondents (93%) reported having no vehicle for transporting waste to the processing site. They therefore evacuated waste on site (Table 4). Most of participants reported using artisanal incinerator in the health facilities were artisanal (42,6%), while more than half reported not having any

**Table 4.** Characteristics of waste management and disposal facilities, Centre Region, March 2024 (*n*=61)

| Variable | Count(n) | Frequency (%) |
| --- | --- | --- |
| Presence of a grinding/disinfection unit | 8 | 13.1 |
| **Processing and disposal** |  |  |
| Out of hospital | 29 | 47.5 |
| In hospital | 32 | 52.5 |
| **Management system used** |  |  |
| Incineration | 48 | 78.7 |
| Landfill | 6 | 9.8 |
| Landfill/incineration | 1 | 1.6 |
| Landfill/incineration/disinfection | 1 | 1.6 |
| Disinfection | 1 | 1.6 |
| On-site Disposal (Discharge) | 3 | 4.9 |
| Disinfection/Discharge (on-site abandonment) | 1 | 1.6 |

We note that 86% of the health facilities visited did not have waste treatment equipment on site. Less than 7% of districts had a conventional incinerator.

Nearly 5% dumped their waste in a public dump. Over 85% used incineration and landfill for disposal (Table 4).

Of the 55 health facilities surveyed, approximately half lacked an incinerator, while only 7.3% possessed a conventional incinerator (Figure 5).

**Fig 5.**
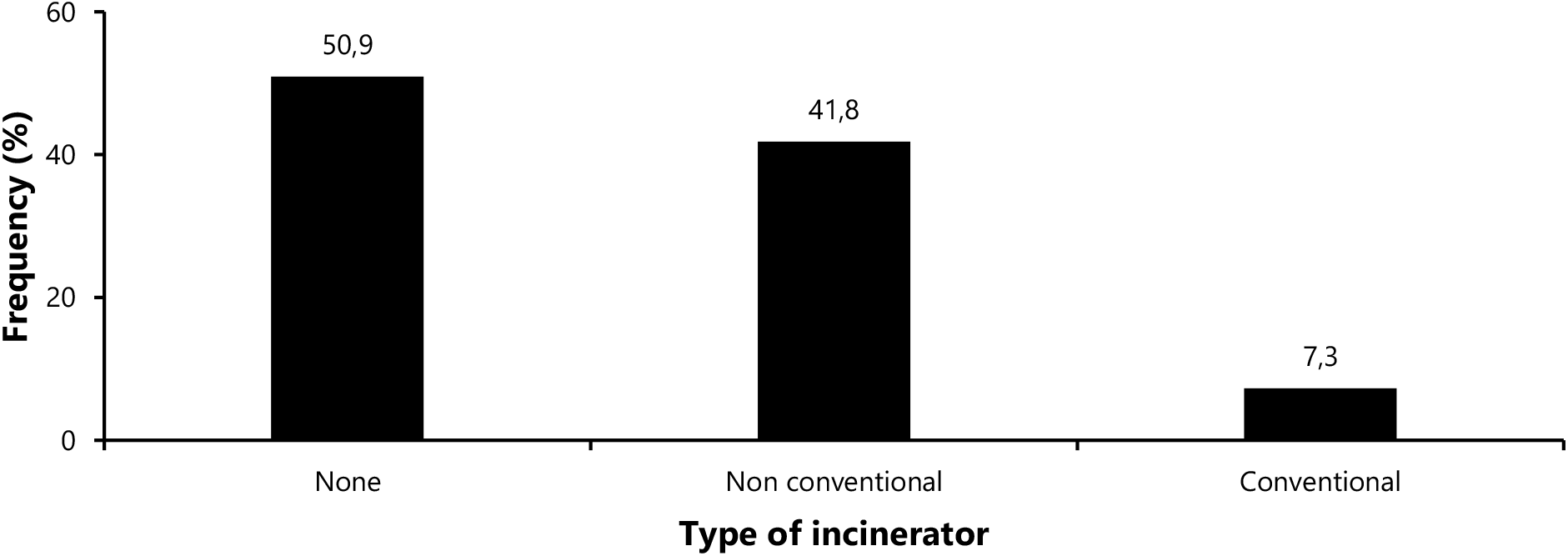
Type of incinerator according to health facility, Centre Region, March 2024 (n=55)

## Discussion

### Socio-professional profile

This study revealed the issue of a lack of qualified hospital waste management personnel, specifically the absence of sanitary engineering technicians, coupled with a dearth of skills among those currently on site. A similar situation pertains with regard to the equipment designated for hospital waste management in the central region. The diversity of personnel observed in this study suggests that a certain level of service provision is being provided in these districts. However, despite the training received by these professionals in the field of hospital hygiene, there is a dearth of sanitary engineering professionals in this human resource. The direct consequence of this is the poor management of hospital waste, which can lead to the proliferation of nosocomial infections, the majority of which are multi-resistant. These findings are consistent with those reported from the North Cameroon region in 2016, which also identified a shortage of both qualitative and quantitative staff. [14]. This can be attributed to a lack of awareness or implementation of Order No. 003/MINEPDED of 15 October 2012, which outlines the requirements for the management of medical and pharmaceutical waste. Article 4 of this order stipulates that any generator of medical and pharmaceutical waste must establish an internal management system comprising a unit responsible for management, qualified and trained staff, a register of activities and adequate equipment. [15]. Furthermore, the unequal allocation of human and material resources within health facilities and health districts contributes to this issue.

### Sorting and packaging

The survey findings indicated that material resources and packaging equipment for hospital waste management are inadequate and do not meet the criteria indicated by the WHO. This is due to the quality of the bags (fragile and lacking labelling), buckets and containers chosen, which do not meet the required standards. A similar situation was observed in the North region, where the majority of health facilities (91.70%) had safety boxes and waste buckets, despite these items being non-compliant. [14]. In accordance with the standards set forth by the World Health Organization (WHO), waste collection bags must adhere to specific criteria. These include being rigid, labelled, tear-resistant, watertight, with a sufficient opening, incinerable, and of a volume that aligns with the quantity of waste generated. This may be attributed to a lack of knowledge among staff regarding hospital waste management norms and standards, as well as a scarcity of material resources, as evidenced by a study conducted in Morocco by El Maaroufi Yacine *et al*. [16]. Therefore, by contextualizing the means and methods of sorting and packaging, such as labels marked and stuck on lidded refuse buckets, which themselves contain refuse bags, this could enable healthcare providers to package waste in a more effective manner while simultaneously categorizing it. The majority of sharps waste was packaged in safety boxes (77.05%). Nevertheless, the packaging of some waste in bin bags elevated the probability of direct injury and exposure to infections such as HIV and viral hepatitis.

### Collection, storage and transport

The intermediate storage of waste in health facilities is an ineffective method, as it is ultimately used as final storage. The prolongation of waste management procedures results in an increase in the proliferation of microorganisms and the generation of unpleasant odors, which are further exacerbated by the effects of climate change. The majority of the waste collected was stored for an undetermined period of time, which in some cases exceeded the legally prescribed period of 48 to 72 hours before being transported. The absence of transportation resources and other measures to guarantee the transfer of waste to treatment facilities, coupled with the scarcity of financial resources or local service providers, results in the accumulation of waste on the premises.

### Treatment and disposal

A variety of techniques are employed in the treatment of hospital waste. Such methods include incineration for infectious healthcare waste. The most prevalent methods of disposal in this study are landfill and incineration. Furthermore, the findings indicate that small-scale incinerators were the most prevalent method of waste disposal.

## Conclusions

The study results highlight a significant shortage of human and material resources to effectively manage hospital waste in most of the health facilities of the Region. To close this gap, it is crucial to provide these facilities with the necessary resources to ensure safe hospital waste management. To achieve this goal, financial resources must be mobilized and secured.

## Data Availability

All data generated or analyzed during this study are included in this published article.

## Declarations

### Authors’ Contribution

Study design & conception: MFE; Data collection: MFE; Data analysis, visualization and interpretation: FZLC; Drafting of original manuscript: MFE; Critical revision of the manuscript: MFE, FZLC, PSDS, FENM, BNS, FKN and TM; Final approval of the manuscript: All authors.

### Ethical Approval Statement

Ethical clearance for the present study was waived by the faculty of medicine ethical review board. Administrative approval was obtained from the Regional Delegate of Public Health for the Centre Region. Additionally, participants were required to give their consent for their responses to be used for research purposes. The confidentiality, anonymity and autonomy of research participants were respected throughout the study. All methods were performed in accordance with relevant guidelines of Helsinki declaration.

### Consent for publication

Not applicable.

## Availability of data and materials

All data generated or analyzed during this study are included in this published article.

## Acknowledgements

We would like to express our gratitude to the healthcare personnel in the health districts outside Yaounde in the Center region who agreed to take part in this study, and to the regional public health delegate for the Center, Dr. AZOUMBOU MEFANT Thérèse, who gave permission for the study to be carried out in her administrative area.

